# Longitudinal COVID-19 Surveillance and Characterization in the Workplace with Public Health and Diagnostic Endpoints

**DOI:** 10.1101/2020.07.25.20160812

**Authors:** Manjula Gunawardana, Jessica Breslin, John M. Cortez, Sofia Rivera, Simon Webster, F. Javier Ibarrondo, Otto O. Yang, Richard B. Pyles, Christina M. Ramirez, Amy P. Adler, Peter A. Anton, Marc M. Baum

## Abstract

**Background:** The rapid spread of severe acute respiratory syndrome coronavirus 2 (SARS-CoV-2) and the associated coronavirus disease 2019 (COVID-19) have precipitated a global pandemic heavily challenging our social behavior, economy, and healthcare infrastructure. Public health practices currently represent the primary interventions for managing the spread of the pandemic. We hypothesized that frequent, longitudinal workplace disease surveillance would represent an effective approach to controlling SARS-CoV-2 transmission among employees and their household members, reducing potential economic consequences and loss of productivity of standard isolation methods, while providing new insights into viral-host dynamics.

**Methodology and Findings:** On March 23, 2020 a clinical study (OCIS-05) was initiated at a small Southern California organization. Results from the first 3 months of the ongoing study are presented here. Study participants (27 employees and 27 household members) consented to provide frequent nasal or oral swab samples that were analyzed by RT-qPCR for SARS-CoV-2 RNA using CDC protocols. Only participants testing negative were allowed to enter the “safe zone” workplace facility. Optional blood samples were collected at baseline and throughout the 3-month study. Serum virus-specific antibody concentrations (IgG, IgM, and IgA) were measured using a selective, sensitive, and quantitative ELISA assay developed in house. A COVID-19 infection model, based on traditional SEIR compartmental models combined with Bayesian non-linear mixed models and modern machine learning, was used to predict the number of employees and household members who would have become infected in the absence of workplace surveillance.

Two study participants were found to be infected by SARS-CoV-2 during the study. One subject, a household member, tested positive clinically by RT-qPCR prior to enrollment and experienced typical COVID-19 symptoms that did not require hospitalization. While on study, the participant was SARS-CoV-2 RNA positive for at least 71 days and had elevated virus-specific antibody concentrations (medians: IgM, 9.83 µg mL^-1^; IgG, 11.5 µg mL^-1^; IgA, 1.29 µg mL^-1^) in serum samples collected at three timepoints. A single, unrelated employee became positive for SARS-CoV-2 RNA over the course of the study, but remained asymptomatic with low associated viral RNA copy numbers. The participant did not have detectable serum IgM and IgG concentrations, and IgA concentrations decayed rapidly (half-life: 1.3 d). The employee was not allowed entry to the safe zone workplace until testing negative three consecutive times over 7 d. No other employees or household members contracted COVID-19 over the course of the study. Our model predicted that under the current prevalence in Los Angeles County without surveillance intervention, up to 7 employees (95% *CI* = 3-10) would have become infected with at most 1 of them requiring hospitalizations and 0 deaths.

**Conclusions:** Our clinical study met its primary objectives by using intense longitudinal testing to provide a safe work environment during the COVID-19 pandemic, and elucidating SARS-CoV-2 dynamics in recovering and asymptomatic participants. The surveillance plan outlined here is scalable and transferrable. The study represents a powerful example on how an innovative public health initiative can be dovetailed with scientific discovery.

## Introduction

The rapid spread of severe acute respiratory syndrome coronavirus 2 (SARS-CoV-2) and the associated coronavirus disease 2019 (COVID-19) have precipitated a global pandemic heavily challenging our social behavior, economy, and healthcare infrastructure. The number of confirmed COVID-19 cases and deaths are rising rapidly. At the time of writing, the U.S. has surpassed 138,000 deaths from the pandemic, with reported global deaths approaching 600,000.

Worldwide, recovery from this active, devastating outbreak cannot begin until safe and effective medicines for treatment and prevention are available. In the interim, it is essential to use the rapidly expanding scientific knowledge on the novel SARS-CoV-2 to update guidelines for COVID-19 patient management as well as protection of the uninfected population. The implementation of social distancing, frequent and thorough hand washing, isolation, and the use of face masks [1] have helped curb the incidence of COVID-19 in many parts of the U.S. Our ability to design and introduce effective public health measures to reduce the spread of COVID-19 relies on an understanding of the viral pathogenesis and dynamics, fundamental aspects of the disease that remain largely unknown.

Workplace SARS-CoV-2 transmission is believed to represent an important contributor to the global COVID-19 pandemic, especially as countries attempt to spur economic activity [2]. In a significant proportion of infected individuals, the disease manifests itself with mild or no symptoms [3] and managing asymptomatic carriers, so-called “silent spreaders”, represents a significant challenge in controlling the pandemic [4]. This concern is heightened by findings suggesting that SARS-CoV-2 transmission rates are similar in symptomatic and asymptomatic infected individuals [5-7]. Workplace transmission from SARS-CoV-2 carriers with mild or no symptoms therefore is a potentially important mode of spreading the highly communicable disease and represents a significant occupational health risk. Infected workers subsequently are susceptible to transmitting the virus further to household members. When workplace COVID-19 acquisition occurs, standard protective measures require all employees who have been in contact with the infected individual to self-quarantine for 2 weeks. This can effectively close units and businesses even though a proportion of the isolated workers may not have contracted the virus. Based on this rationale, the development and assessment of measures to effectively control SARS-CoV-2 transmission in the workplace is highly significant and necessary to provide a safe occupational environment.

In a May 15, 2020 interview, Dr. James W. Curran, Dean, Rollins School of Public Health at Emory University, stated, “Accurate surveillance is the conscience and guidepost for public health” [8]. The current clinical study builds on this foundational principle and consists of longitudinal and intensive characterization of COVID-19 prevalence and incidence at the Oak Crest Institute of Science (Oak Crest), a nonprofit science research organization in Southern California, USA. The two primary study objectives were to: (1) characterize the rate of COVID-19 acquisition in a small cohort of workers interacting on a daily basis in the workplace; and (2) determine the utility of these data in managing workplace COVID-19 exposure, minimizing further spread as per public health advisories. Exploratory aims include characterizing SARS-CoV-2 transmission between Oak Crest employees and their household members and measuring the viral dynamics in infected individuals from the study cohort.

## Methods and Materials

### Ethics Statement

All human research under OCIS-05, “Longitudinal Characterization of COVID-19 Prevalence and Incidence in a Small Working Institution with Both Public Health and Diagnostic Aims”, was approved by Aspire IRB (Aspire Study # 1281548) and conducted according to the Declaration of Helsinki. All 54 study participants provided written informed consent or assent.

Because of the nature of the study population, participants were informed in the consent form that, “Although all efforts will be made to protect your privacy and confidentiality (we will explain more below), other people in your house and at OCIS will know you are in the study. The results of your tests could become known if you have to be quarantined at home.”

### Clinical Study Design

The clinical study was initiated on March 23, 2020 and is ongoing at the time of writing. Results from the first three months of testing (March 23-June 22, 2020) are presented here. All employees, students, and volunteers at Oak Crest (oak-crest.org), a small nonprofit academic science research organization located in Monrovia, CA, USA, were asked to participate in the prospective, longitudinal, observational study designed to last 12 weeks, or longer. Those choosing not to participate had no negative employment or finance-related consequences, but were asked to work from home exclusively. Household members from the above study population also were invited to participate in the study. At study onset, up to 30 Oak Crest employees and up to 60 household members were anticipated to participate.

Volunteers who met the inclusion criteria were asked to provide written consent (or assent in the case of minors) to participate in the study, under an institutional review board approved protocol. At baseline, participants also completed a survey that included basic demographic information (name, address, phone number, date of birth, gender, race, and ethnicity), relevant co-morbid conditions, contact with possible COVID-19 positive individuals, and recent travel history (**Table 1**). A symptom diary also was completed (**Table 1**). Where relevant, entries were updated weekly. The above items were the extent of personal history and demographics collected. No medical records were obtained/reviewed.

**Table 1.**
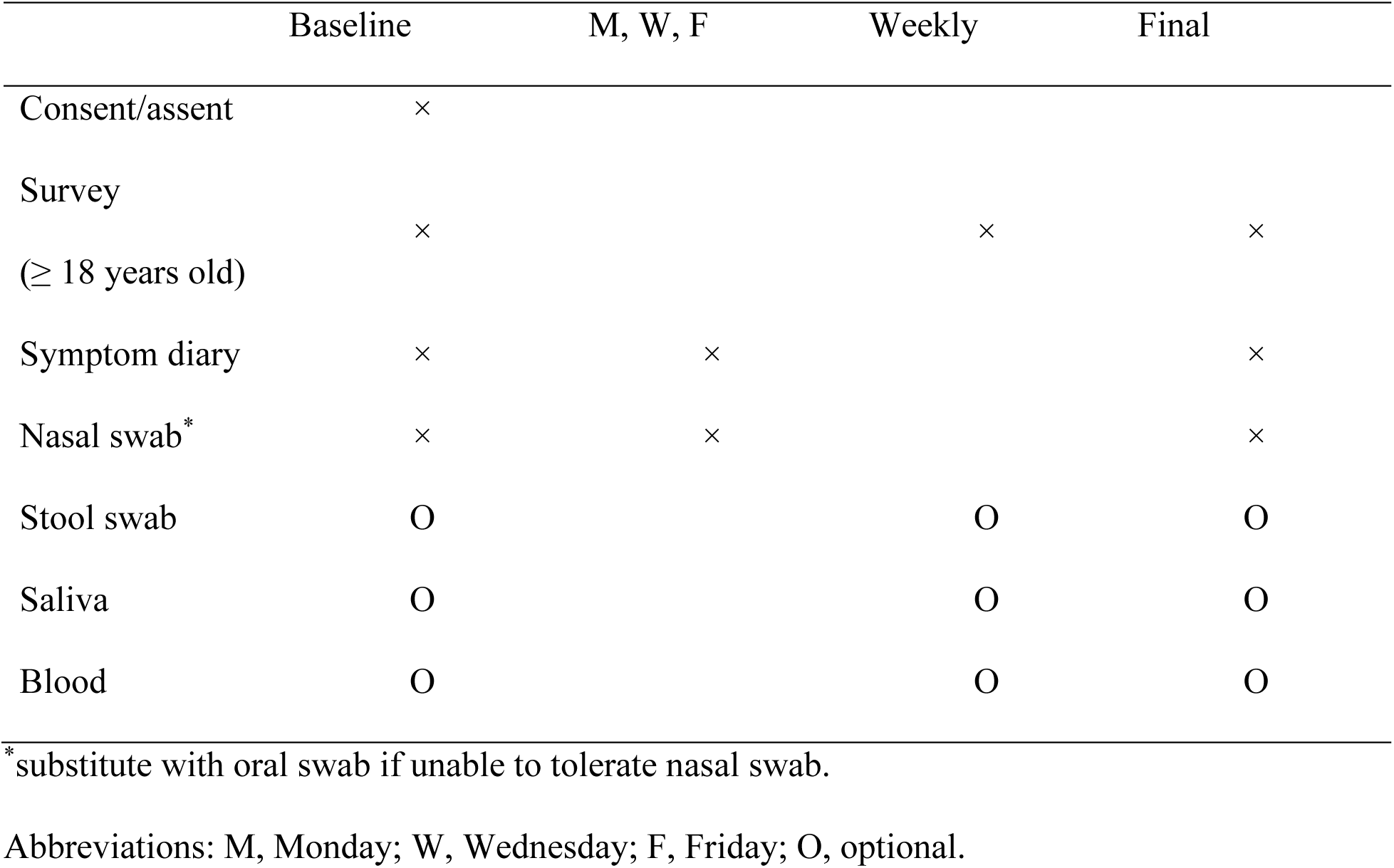
Study evaluations.

The clinical study was unblinded and there were no study-specific interventions. Descriptive and summary statistics are provided for this small observational study.

Test results for nasal or oral swabs collected in the morning typically were available early in the afternoon of the same day. When a participant tested positive or the results were inconclusive, testing was repeated for confirmation or collected at increased frequency (daily when feasible) until the participant had repeated negative results. Participants were advised in the informed consent and, again, when informed of research test results that these results were research-based and not intended for clinical diagnosis. Participants received referrals for testing and recommendations (e.g., self-quarantine) based on the current, preferred public health requirements at the time.

### Sample Collection

Nasal sample collection was overseen by two researchers equipped with personal protective equipment (PPE) in the Oak Crest parking lot, starting at 8:30 AM. Study participants were required to wear masks, waited in their parked vehicles, and received a sterile nasopharyngeal swab (synthetic applicator tip, synthetic handle) from one of the two study researchers (**Fig 1A**).

**Figure 1.**
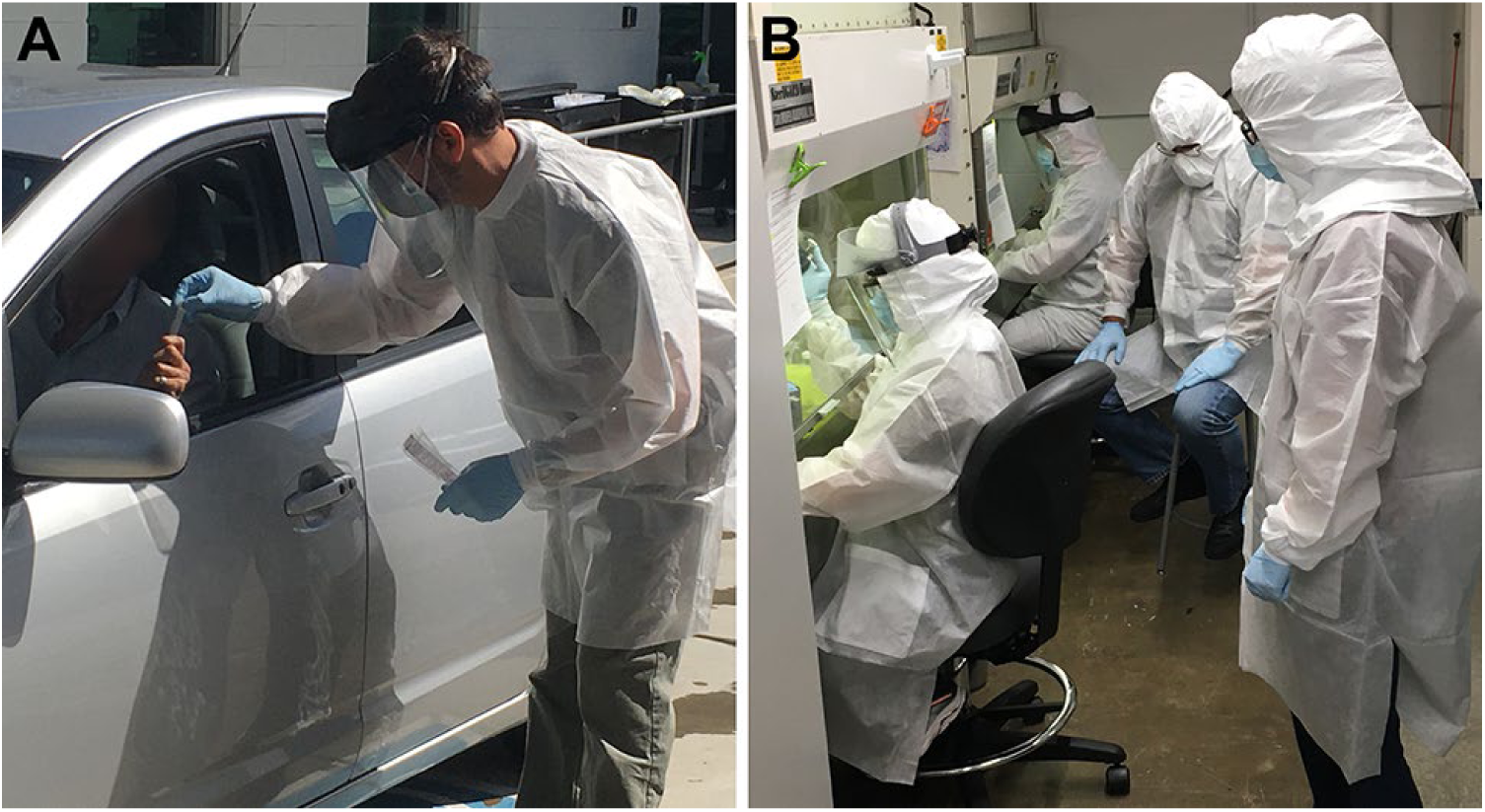
Photographs documenting sample collected and analysis in the OCIS-05 clinical study. (**A**) Sample collection from participant who previously self-collected a nasal swab specimen the participant’s vehicle. (**B**) Extraction of SARS-CoV-2 RNA by study team.

Due to limited availability, five different swab types/lots were used over the course of the study, in chronological order: (1) eSwab (Model 220246, BD Diagnostics, Sparks, MD); (2) sterile polyester tipped applicator (Model 25-806 2PD, Puritan Medical Products, Guilford, ME); (3) flocked collection device (Model 25-3406-H, Lot 2937, Puritan Medical Products); (4) flocked collection device (Model 25-3000-H, Puritan Medical Products); and (5) flocked collection device (Model 25-3406-H, Lot 7221, Puritan Medical Products). Prior to sample collection, the subject closed the vehicle window and self-collected one nasal specimen (one swab, both nares) according to the study instructions. An oral swab sample (gums, cheeks, and back of throat) was collected according to the study instructions *in lieu* of the nasal swab if the latter could not be tolerated. The used swab tip was placed in a pre-labeled, empty (i.e., dry) microfuge tube (1.5 mL), the swab handle broken off, and the closed tube handed to the researcher *via* the open window. If the participant was uncomfortable breaking the swab handle, the used swab was handed to the researcher who then broke it and sealed the tip in the sample tube. The sample tube was sprayed with 70% v/v isopropanol and placed on wet ice until all samples were collected. Typically, 30 samples or fewer were collected in under 75 min. Under certain circumstances, study participants self-collected swab samples at home and placed the swab tip in a microfuge tube (1.5 mL) containing RNA Shield (300 µL, Zymo Research, Tustin, CA). The samples were stored and transported at room temperature or 4°C and processed within 60 h. Control experiments showed that RNA integrity was maintained under these conditions.

Optional saliva samples were self-collected in Falcon tubes (50 mL) at the participant’s home or in their sealed vehicle and stool swabs were collected at the participant’s home. Specific written instructions were provided to participants opting to provide these specimens. Blood (5-8 mL, ×2) using Vacutainer® (Becton, Dickinson and Company, Franklin Lakes, NJ) tubes for serum (spray-coated silica) and plasma (spray-coated K_2_EDTA) was collected by a licensed phlebotomist in the Oak Crest parking lot, while the participant remained comfortably seated in their vehicle.

### Chemicals and Reagents

All reagents were obtained from Sigma-Aldrich (St. Louis, MO), unless otherwise noted.

### RNA Extraction

RNA was extracted from swab specimens in the collection tubes using the Quick RNA Viral Kit (R1036, Zymo Research) according to the manufacturer’s instructions consistent with the CDC COVID-19 testing kit guidelines [9].

### RT-qPCR Analysis

One-step reverse transcription (RT) and quantitative PCR (qPCR) reaction was performed using the TaqPath™ 1-Step RT-qPCR Master Mix (ThermoFisher Scientific, Waltham, MA) in a 20 µL final reaction volume per the manufacturer’s instructions. Three target sequences were amplified simultaneously [10] in accordance with CDC diagnostic COVID-19 testing guidelines [9]: two SARS-CoV-2 nucleocapsid protein (*N*) gene transcript fragments (N1 and N2); and one human RNase P (*RP*) gene transcript fragment (RP). The primer/probe sequences are as described elsewhere [9] and were obtained from Integrated DNA Technologies (10006606, 10006713, Coralville, IA). RNA standards for SARS-CoV-2 nucleocapsid (*N*) was obtained from Microbiologics (HE0060-100-T, St. Cloud, MN) and served as positive controls. Confluent Caco-2 cells were used as a human specimen control (HSC) according to CDC guidelines [9].

Every panel consisted of one 96-well plate that accommodated 28 clinical specimens, each with three PCR probes, (28×3 = 84 wells), HSC (1×3 = 3 wells), SARS-CoV-2 RNA standard as a positive control (1×3 = 3 wells), a no template control (1×3 = 3 wells), leaving three wells that were used for additional validation (e.g., naïve swab). This panel configuration meets or exceeds the CDC guidelines [9].

96-well PCR plates were prepared using reagents on ice and centrifuged at 1,000 RPM for 1 min at room temperature. The plates were run on a CFX96 Touch Real-Time PCR Detection System (Bio-Rad Laboratories, Inc., Hercules, CA) using the following cycling conditions: Stage 1, 25°C, 2 min (1 cycle); Stage 2, 50°C, 15 min (1 cycle); Stage 3, 95°C, 2 min (1 cycle); Stage 4, Step 1, 95°C, 3 s, Step 2, 55°C, 30 s; Stage 4, loop of 45 cycles of Step 1 and 2 (total 46 cycles per assay). Stages 1-2, reverse transcription; Stage 3, inactivation; Stage 4, amplification. The normalized SARS-CoV-2 RNA copy number, Δ*C*_*t*_ was calculated according to eq. 1:

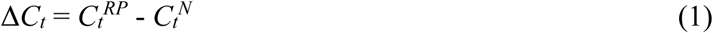

where *C*_*t*_^*RP*^ is the *C*_*t*_ value for the *RP* gene transcript in the sample and *C*_*t*_^*N*^ is the mean *C*_*t*_ for corresponding two SARS-CoV-2 *N* gene transcripts (N1 and N2).

### SARS-CoV-2 *N* Gene RT-qPCR Probe Calibration

Calibration plots of *Ct versus* RNA copy number were generated for every batch of TaqPath™ 1-Step RT-qPCR Master Mix kits. Predetermined amounts of RNA standard for SARS-CoV-2 *N* gene transcript fragments (*vide supra*) were serially diluted in nuclease-free water (Promega Corporation, Madison, WI). In a typical experiment, eight calibration standards were used, spanning the 0.15-1.5×10^6^ RNA copy number range. The samples (20 µL) were analyzed (N1 and N2 probes) as described above.

### Assay Result Interpretation

The diagnostic panel result interpretation followed CDC guidelines [9]. Specimens with a cycle threshold (*Ct*) less than 40 for N1, N2, and RP were considered positive. Samples with a *Ct* greater than 40 for N1 and N2, but less than 40 for RP were considered negative. Samples with a *Ct* for either N1 or N2 less than 40 (i.e., only one of the two probes resulted in a *Ct* less than 40) and a *Ct* less than 40 for RP were considered inconclusive. Samples with a *Ct* for RP greater than 40 were considered invalid.

### Swab Assessment

A range of collection swabs were used in the clinical study due to supply shortages (see **Results** section). The two most common swab types employed were model 25-2000-H and 25-806-2PD (Puritan, Guilford, MA). All swabs were evaluated for fungal and bacterial contamination by qPCR using published methods [11, 12]. Swab batches that were found to be free of microbial contamination were evaluated for nasal sample collection efficiency. Typically, 3-6 volunteers from the clinical study self-swabbed nasally according to the study protocol with the test swabs. The swabs were analyzed as described above using the RP probe, and *Ct* values below 25 typically were indicative of efficient sample collection. Swabs from that batch then were used in the study.

### Quantification of Serum IgG, IgM, and IgA against SARS-CoV-2

#### Soluble SARS-CoV-2 Spike Receptor Binding Domain Protein Production

SARS-CoV-2 spike receptor binding domain (RBD) was produced by transient transfection of HEK-293F cells with the plasmid pCAGGS [13, 14] containing the SARS-CoV-2, Wuhan-Hu-1 Spike Glycoprotein Gene RBD with C-Terminal Hexa-Histidine Tag (generously provided by Dr. F. Krammer).

Transfection was carried utilizing the Expi293™ Expression System (Gibco, ThermoFisher Scientific) following the manufacturer’s instructions. RBD was purified by affinity chromatography utilizing Ni-NTA agarose (ThermoFisher Scientific) and RBD purity was assessed by SDS-PAGE.

#### Enzyme-linked Immunosorbent Assay (ELISA) for Antibodies against RBD

Antibody responses against RBD were measured in a modified ELISA based on a protocol by Krammer *et al*. [13, 14]. In brief, 96-well microtiter plates were coated with soluble RBD protein (2 µg mL^-1^) in calcium- and magnesium-free phosphate buffered saline (PBS, Gibco). The plates were washed three times in PBS containing 0.1% Tween-20 (TPBS) and incubated with PBS containing 3% dried milk (bioWORLD, Dublin, OH) for a minimum of one hour at room temperature before removal. Participant serum was added in duplicates in three-fold serial dilutions from 1:40 to 1:1080 in PBS and incubated at room temperature for two hours. The plates were washed three times with TPBS, and the secondary antibody anti-human IgG-horseradish peroxidase (Bethyl Laboratories, Montgomery, TX) –or anti-human IgM-horseradish peroxidase, or anti-IgA-horseradish peroxidase– was added in PBS at a 1:50,000 dilution for incubation at room temperature for one hour. The plates were washed three times with TPBS, followed by addition of 100 µL TMB substrate solution (ThermoFisher Scientific) for 10 minutes at room temperature and then 100 µL sulfuric acid stop solution (ThermoFisher Scientific). The plates were read at 450 and 650 nm wavelengths on a Spark 10M microplate reader (Tecan, Baldwin Park, CA). Each plate also contained wells with the control anti-RBD monoclonal IgG antibody CR3022 (Creative Biolabs, New York, NY) plated in serial dilutions to establish a standard curve, or an IgM or IgA monoclonal antibody with the same variable region as CR3022 produced in-house.

Briefly, IgA and IgM RBD-specific control antibodies were produced by transfection of 293F cells with plasmids coding the CR3022 light chain, the J chain and the human IgA or IgM heavy chains containing the heavy variable region of CR30222 (plasmids were kindly provided by G. Alter). Five days after transfection, antibodies were purified by affinity chromatography. Purity and antibodies chain stoichiometries were assessed by SDS-PGE under reducing and non-reducing conditions.

Optical density values from participant serum were compared to the standard curve to extrapolate equivalence to a concentration of the control IgG, IgM, or IgA antibody.

### Model Analysis

A COVID-19 infection model that combines traditional SEIR compartmental models, along with Bayesian case velocity and machine learning was used for model analysis [15]. This model fuses three methods to provide accurate predictions of case counts and hospitalizations. Briefly, a Bayesian nonlinear mixed model was fitted to the velocity (first-derivative) of the cumulative case counts for each location, such as California, or a specific county, zip code, or even workplace and incorporated prior information such as local interventions to obtain the posterior distribution of the trajectory of the cases. This was then fed into the compartmental model to predict deaths using the random forest algorithm trained on COVID-19 data and population-level characteristics such as age, expected proportion of co-morbidities, gender, and race/ethnicity. This yielded daily projections and interval estimates for cases and deaths for each location. The approach combined the strengths of traditional epidemiologic compartmental models with curve-fitting statistical models and modern machine learning.

### Data Analysis

Datasets were analyzed using GraphPad Prism (version 8.4.3, GraphPad Software, Inc., La Jolla, CA). Statistical significance was defined as two-sided *P* < 0.05. The unpaired, parametric, two-tailed *t* test was used to compare nasal and oral swab *RP* gene transcript *Ct* values.

The probability of multiple, sequential false negative SARS-CoV-2 results was estimated using traditional probability theory. Each serial test was assumed to be independent as the probability of a test result outcome was not dependent on the result of the previous test. The sensitivity of the test was assumed to be 0.99 and the specificity 0.95. The probability of being COVD-19 positive given a negative test result was estimated according to Bayes’ theorem, eq. 2:

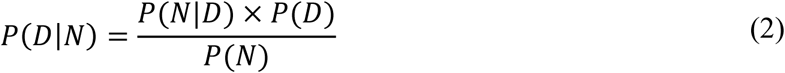

where *P*(*D*|*N*) is a conditional probability, the likelihood of event *D* occurring given that *N* is true; *P*(*N*|*D*) is a conditional probability, the likelihood of event *N* occurring given that *D* is true; *P*(*D*) and *P*(*N*) are the probabilities of observing *D* and *N*, respectively. This probability was estimated for prevalence ranges from 0.05 to 0.5 and the greatest upper bound was taken as the probability bound.

## Results

### Study Participants

A total of 54 subjects have participated in the study over the first three months, 27 Oak Crest employees, students, and volunteers and 27 household members. The corresponding demographics are presented in **Table 2**.

**Table 2.** Demographics of study participants.

|  | Oak Crest Employees | Household Members |
| --- | --- | --- |
| Participants enrolled, *n* | 27 | 27 |
| Age (years), median (range) | 39 (21-68) | 40 (11-80) |
| Female, *n* (%) | 13 (48) | 16 (59) |
| Male, *n* (%) | 14 (52) | 11 (41) |
| Race and Ethnicity, *n* (%) |  |  |
| Black or African American | 0 | 0 |
| White | 23 (85) | 25 (93) |
| Hispanic | 10 (37) | 14 (52) |
| Non-Hispanic | 13 (48) | 11 (41) |
| Asian | 2 (7) | 2 (7) |
| Other | 2 (7) | 0 |

### Sample Collection Efficiency

The majority of clinical specimens consisted of nasal swab samples. Only 3 of the 54 participants (5.6%) could not tolerate the nasal swab procedure and self-collected oral samples instead. A total of 942 samples were analyzed in the three-month study. **Figure 2A** compares the distribution of *RP* gene transcript *Ct* values (i.e., indication of host RNA recovered) for the nasal (*N* = 831; 88.2%) and oral (*N* = 111; 11.8%) samples. The median (IQR) values were: nasal swab, 23.3 (22.3-24.4); oral swab, 25.1 (24.6-26.1). The 2 populations were significantly different (*P* < 0.0001) with lower observed median *Ct* values (i.e., more host RNA collected) for the nasal swab group.

**Figure 2.**
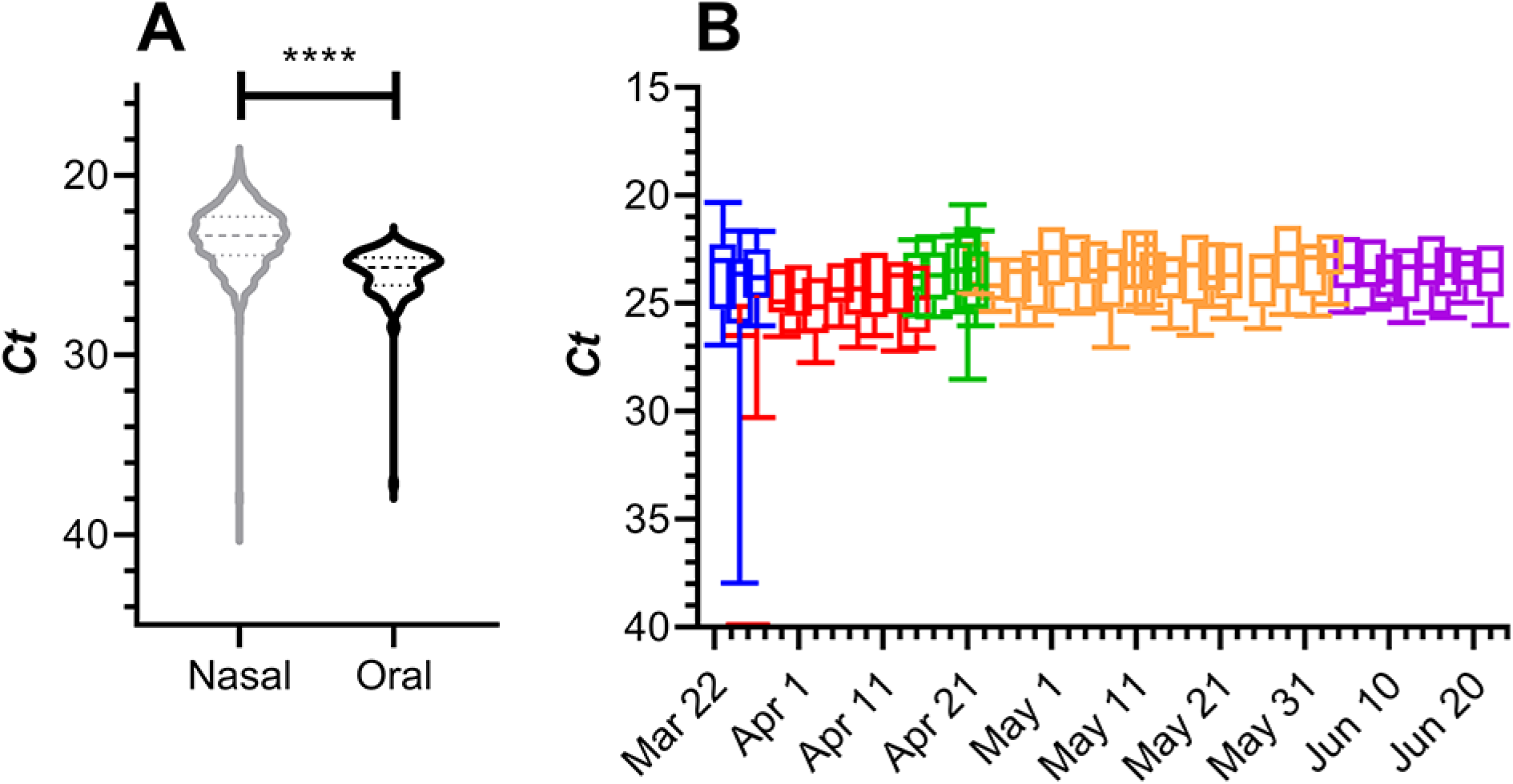
Distribution of *RP* gene transcript *Ct* values over the three-month clinical study. **(A)** Violin plot comparing the *Ct* value for nasal (grey) and oral (black) swab samples. Broken horizontal lines represent medians (dashed) and quartiles (dotted). Comparison of the nasal and oral swab group *Ct* values using an unpaired, parametric, two-tailed *t* test showed that the groups were significantly different (*P* < 0.0001). **(B)** Box plots of *Ct* values from nasal and oral swab samples for each study day. The box extends from the 25th to 75th percentiles, with the horizontal line in the box representing the median; whiskers represent the 10-90^th^ percentiles. Colors are representative of different nasopharyngeal swab types: blue, Model 220246, BD Diagnostics; red, Model 25-806 2PD, Puritan Medical Products; green, Model 25-3406-H, Lot 2937, Puritan Medical Products; orange, Model 25-3000-H, Puritan Medical Products; magenta, Model 25-3406-H, Lot 7221, Puritan Medical Products.

Due to a shortage of appropriate swabs at the onset of the study, 5 different swab types –each tested prior to use as described in the **Methods and Materials** section– were used. **Figure 2B** compares the distribution of *RP* gene transcript *Ct* values over the 3-month study as a function of swab type. Median *Ct* values consistently were below 25, suggesting efficient sample collection.

### RT-qPCR Measurement Reliability

Analysis of data from the study did not reveal any demonstrated false negative or false positive results from the RT-qPCR measurements. The lower limit of detection of the SARS-CoV-2 *N* gene transcript fragment RT-qPCR assay was 10 RNA copies per reaction. All positive results were confirmed by re-testing on multiple, successive occasions (*vide infra*). Any false negative results would need to have been in asymptomatic individuals who did not infect other employees or household members. A probabilistic analysis on an employee with 12 consecutive false negative results afforded a *P*-value < 0.000000001 (P = 10^−24^), assuming a 99% sensitivity and 95% specificity. For a participant to have 3 consecutive false negative results calculated over a wide range of prevalence values (0.005-0.5), *P* < 0.0000114.

There were 5 invalid measurements in the first three study days attributed to lack of familiarity with the sampling protocol and therefore these were not included in the analysis. An additional 6 invalid samples were obtained during the remaining study period, accounting for 0.63% [6/(6+948)] of total measurements.

There were a total of 3 inconclusive measurements (0.32%) over the course of the study, with the following caveats: the inconclusive results did not include those obtained for the 2 participants who tested positive for SARS-CoV-2 RNA (*vide infra*), nor did they include 9 inconclusive results obtained on 2 study days (June 3 and 5) for participants who repeatedly tested negative for SARS-CoV-2 RNA. The high number of inconclusive measurements obtained on those 2 consecutive sampling days were attributed to reagent trace contaminants that resulted in erroneous, high *Ct* value (i.e., low RNA copy number equivalent) background noise for amplicons obtained with the N1 or N2 primers.

### Positive Participant RT-qPCR Test Results

Two participants were found to be positive for SARS-CoV-2 RNA over the course of the study (**Fig. 3**).

**Figure 3.**
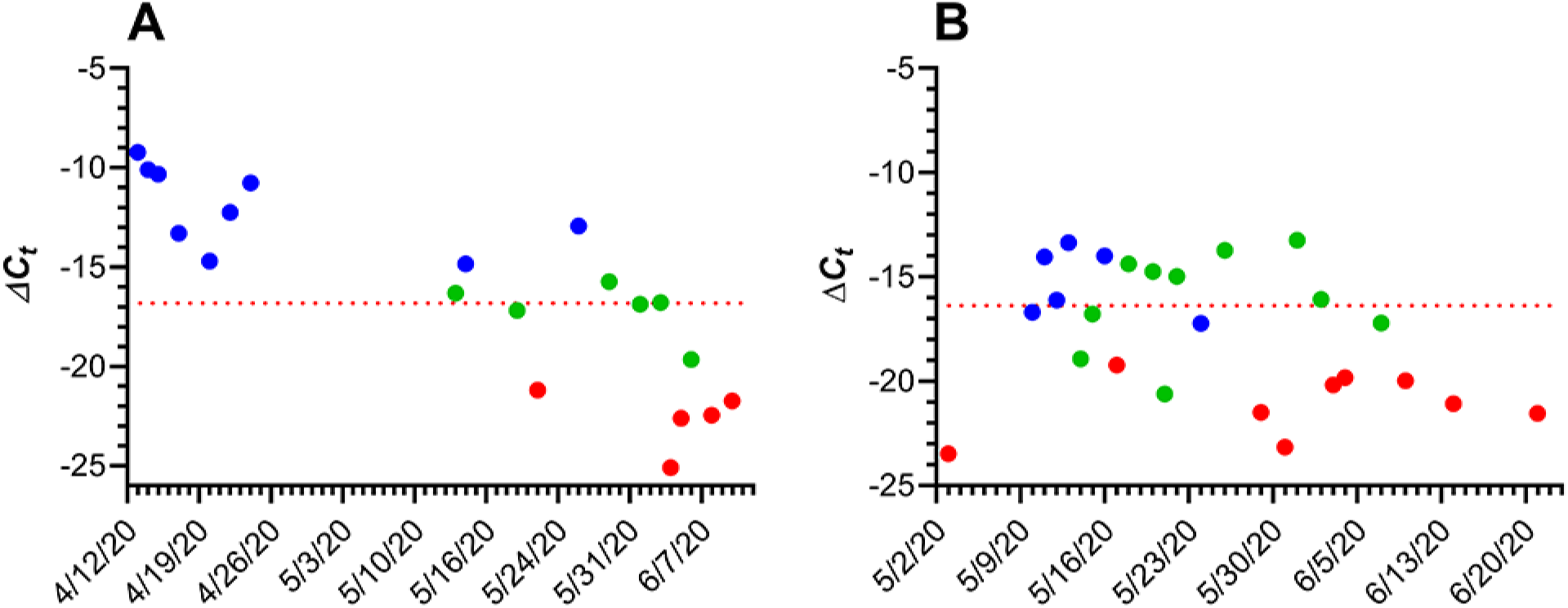
Longitudinal Δ*C*_*t*_ values for two subjects testing positive for SARS-CoV-2 RNA (nasal swabs) during the course of the 3-month study. Blue, positive; green, inconclusive; red, negative. In cases when the measured fluorescent signal for N1 and/or N2 primers did not exceed the background level at the end of the run, the *Ct* value was set to 46. The horizontal red broken line represents the average boundary between positive (higher Δ*C*_*t*_ values) and negative (lower Δ*C*_*t*_ values) based on the mean *RP* gene transcript *Ct* for the subject over that timeframe and N1, N2 primer *Ct* values of 40. (**A**) RT-qPCR results from subject 39, a recovering clinically COVID-19-diagnosed household member; horizontal line at Δ*C*_*t*_ = -16.83; (**B**) RT-qPCR results from subject 31, a COVID-19 asymptomatic employee, horizontal line at Δ*C*_*t*_ = -16.43.

In the participant’s clinical diary, subject 39 (age, over 50 years; further demographics not disclosed for the purposes of confidentiality) described potential SARS-CoV-2 exposure on March 12, with the onset of COVID-19 symptoms on March 16 that persisted until March 31. During that period, the participant was self-isolated without hospitalization and, on April 1, tested positive for SARS-CoV-2 by a clinical RT-qPCR test (oral swab). On April 11, the participant enrolled in our clinical study. The last nasal swab test that met the CDC criteria for a positive result was on May 26, indicating that the participant was COVID-19 positive for at least 71 d (March 16-May 26). Oral (April 14) and stool (April 14, 15) samples were negative for SARS-CoV-2 RNA.

Subject 31 (age, over 50 years; further demographics not disclosed for the purposes of confidentiality) is an employee who only accessed the facility once per week and therefore was tested weekly. The test results went from negative (up to May 3) to positive for SARS-CoV-2 RNA (May 10) and remained positive for 14 d (until May 24). The participant was asymptomatic during that period. Saliva (May 12, 13, 14, 15) and stool (May 13, 14, 15, 16, 17, 18) samples were either negative or inconclusive for SARS-CoV-2 RNA.

### Serological Test Results

A total of 33 serum samples collected between March 27 and May 26, 2020 were analyzed for virus-specific IgM, IgG, and IgA concentrations as described above. The results are summarized in **Table 3** and **Fig. 4**. Other than subject 39, only one participant had detectable serum IgG concentrations (collected on May 15, 0.12 µg mL^-1^).

**Table 3.** Summary of receptor binding domain (RBD) antibody concentrations in participant serum samples at baseline, Mar. 27-Apr. 3, 2020 (*N* = 16) and for subject 39. Because of the high antibody concentrations measured in multiple samples, the data for subject 39 are presented separately.

| | IgM, $\mu\text{g mL}^{-1}$ | IgG, $\mu\text{g mL}^{-1}$ | IgA, $\mu\text{g mL}^{-1}$ |
| --- | --- | --- | --- |
| Limit of Detection (LLD)* | 0.0049 | 0.0017 | 0.0017 |
| % > LLD** | 56 | 0 | 6 |
| Median (IQR), baseline† | 0.39 (0.24-0.96) | N/A | 0.49 |
| Median (range), Subject 39 | 9.83 (2.12-11.4) | 11.5 (11.5-11.7) | 1.29 (0.34-1.70) |

\*Assay limit of detection (LLD) determined at 1:100, 1:40, and 1:20 dilution. The LLD is presented as the diluted concentration.
\*\*Data represent proportions of samples that contained detectable antibody concentrations.
†Interquartile range (25th to 75th percentile).

**Figure 4.**
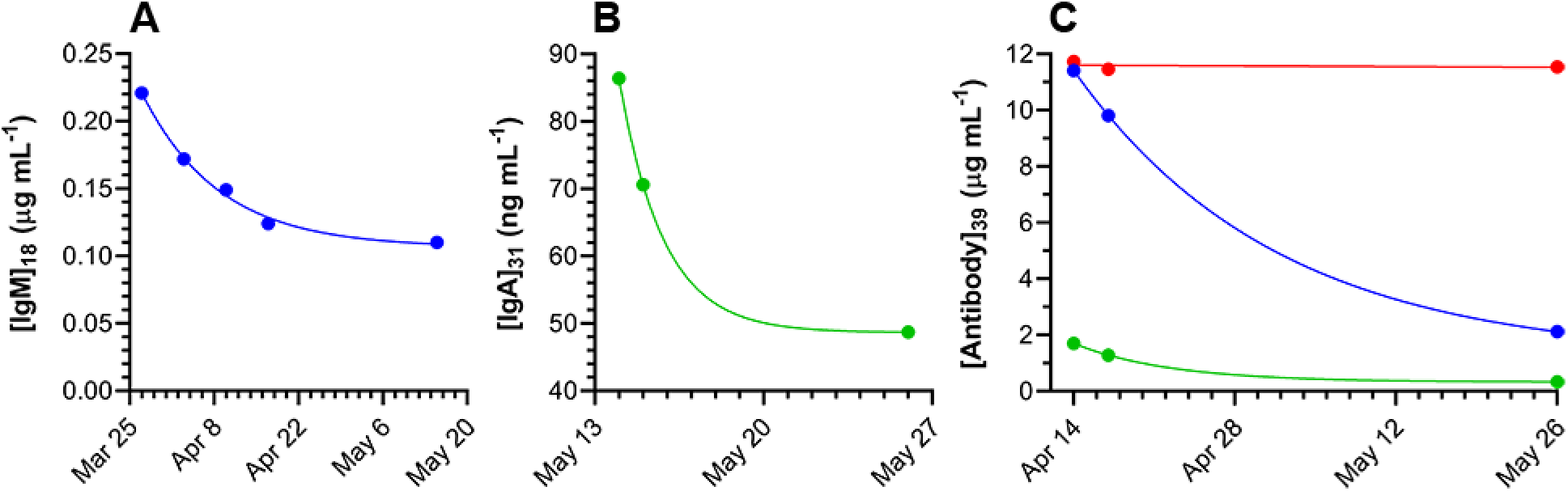
Longitudinal receptor binding domain (RBD) serum antibody concentrations for three study participants; blue circles, IgM; red circles, IgG; green circles, IgA. (**A**) Serum IgM concentrations for subject 18, an employee who experienced symptoms consistent with COVID-19 that resolved just prior to the start of the study, but tested negative for SARS-CoV-2 RNA in nasal swab samples. Blue line, one-phase exponential decay least squares fit (*R*^*2*^ = 0.995) with a half-life of 8.8 d. (**B**) Serum IgA concentrations in subject 31, an employee who was asymptomatic, but tested positive for SARS-CoV-2 RNA in nasal swab samples. Green line, one-phase exponential decay least squares fit (*R*^*2*^ = 1.000) with a half-life of 1.3 d. (**C**) Serum antibody concentrations for subject 39, a household member recovering from clinically diagnosed COVID-19. Red line (IgG), linear regression (*y* = -0.0125*x* + 11.61; *R*^*2*^ = 0.094). Blue (IgM) and green (IgA) lines, one-phase exponential decay least squares fit (*R*^*2*^ = 1.000) with half-lives of 12.3 and 5.8 d, respectively.

### Model Results of Local Study Impacts

The model was run for California with prevalence levels seen on March 23, 2020 and on July 7, 2020 in Los Angeles County. Given 27 employees and 27 household contacts with the characteristics of participants from the current study and without surveillance interventions, the model produced an expectation that up to 7 employees (95% *CI* = 3-10) would have become infected with at most 1 of them requiring hospitalizations and 0 deaths. The lack of deaths stems from the population demographic tending towards younger persons and improved in-hospital treatment procedures at the time of writing.

### Practical Study Considerations

A number of policies and procedures were instituted during the study, as described below. Some were part of the original protocol, and others were added based on practical considerations. While working on site at Oak Crest, all individuals practiced social distancing, hand hygiene, and wore face coverings when in close proximity to other employees on Mondays (*vide infra*); testing negated the need for mandatory face masks on other days. Outside of work, employees and their household members were encouraged to strictly adhere to current public health advisories regarding gathering in groups, wearing face coverings, and social distancing.

Participants who tested positive for SARS-CoV-2 RNA received referrals for testing and recommendations (e.g., self-quarantine) based on the current, preferred public health requirements. Symptomatic participants, even if testing negative for SARS-CoV-2 RNA by RT-qPCR, would have been asked to return home (none were reported in the study), self-quarantine at a minimum until symptoms resolved, and contact their health care provider about confirmatory clinical testing and symptom management. Participants testing positive for SARS-CoV-2, even if asymptomatic, were informed as soon as possible and asked to go home, self-quarantine, and contact their health care provider or the Department of Public Health (DPH) about confirmatory testing, treatment, and quarantine recommendations. Participants also were given educational information to guide other household members of potential risk and some of their options. Any employee who was asked to self-quarantine, or did not choose to participate in the study, worked remotely as much as possible and continued to receive full pay.

Participants who tested negative and obtained a subsequent invalid test result were asked to wear protective face covering (surgical masks were provided by Oak Crest if needed) until they could be re-tested. Initially, participants who tested negative and obtained a subsequent inconclusive test result were sent home immediately and told to self-quarantine until they could be re-tested (i.e., new swab sample collected and analyzed). As the study progressed, that protocol was modified to be the same as an invalid result (i.e., wear protective face covering rather than send the employee home for self-quarantine), assuming that previous test results were negative. The longest period between tests was Friday to the following Monday (3 d). Starting in May, all employees were required to wear protective masks on Monday mornings until the test results were finalized and communicated. The same procedure was instituted for 3-day week-ends when there was an additional day between tests. Employees who missed one day of testing were required to wear a protective mask until they were re-tested. Employees who missed more than one day of testing only were permitted to re-enter the facility after a new sample tested negative.

## Discussion

The current, ongoing clinical study has two primary aims: (1) characterize the rate of SARS-CoV-2 acquisition in a small cohort of participants interacting on a daily basis in the workplace; (2) determine the ability of these data to manage workplace SARS-CoV-2 exposure and consequences, minimizing further spread as per public health advisories. Workplace study participants include staff, students, and volunteer researchers, referred to as “employees”. Family members and housemates also were recruited leading to a sub-cohort referred to as “household members”. Related exploratory study goals include: characterizing the rate of SARS-CoV-2 acquisition in employee and household members; quantifying antibody-specific responses in blood at baseline (previously exposed) and while on study (to capture asymptomatic/pre-symptomatic, newly infected); characterizing viral shedding parameters in saliva and stool samples.

Clinical SARS-CoV-2 RNA test kits that have received Emergency Use Authorization (EUA) by the FDA and are processed in a laboratory certified according to the Clinical Laboratory Improvement Amendments (CLIA). Such kits have been a rare resource during this clinical study and, consequently, were not available for investigative purposes. Our clinical study used research reagents to achieve outcomes, but did not interfere with the availability of reagents in short supply for clinical diagnosis. The RT and amplification kit (TaqPath™ 1-Step RT-qPCR Master Mix) and primers have not been in short supply. The employed RNA extraction kit (Quick RNA Viral Kit) did not have FDA EUA in March 2020 and has been readily available since then. Nasopharyngeal swabs were in short supply early in the study requiring the use of existing, in-house stocks until they became more readily available starting in May 2020 (**Fig. 2B**). The various swab types performed similarly and met the study criteria. To date, we have analyzed approximately 1,000 samples over the course of the 3-month study, compared to over 37.2 million clinical tests reported for the U.S. over a similar period according to the Centers for Disease Control and Prevention [16].

The study was designed to be efficient in terms of manpower commitment and protecting the study team from exposure to SARS-CoV-2 during sampling. Employees were trained in self-sample collection while isolated in their vehicles 3 times per week (Mon, Wed, Fri) between 8:30 and 9:15 AM with support from the study team. The samples (typically 23-28 per day) were processed the same day with results were available by early afternoon. A team of 4 researchers (2 handling samples and 2 observing for quality assurance, **Fig. 1B**) performed the RNA extraction and loading of the 96-well plate for RT and cDNA gene fragment amplification, typically completed within 90 min. Additional labor included quality assurance and subsequent data analysis. These resource commitments made the study feasible to continue indefinitely until recovery from the COVID-19 pandemic allows for a safe workplace.

During the course of the study, one employee (subject 31) became SARS-CoV-2 RNA positive for ca. 2 weeks (**Fig. 3B**). The viral RNA copy number (median, 13.1 copies per swab; IQR, 10.9-18.8 copies per swab) remained low throughout and the subject did not report any COVID-19 symptoms. Blood draws on May 14, 15, and 26 from the subject produced serum samples that were below the analytical LLD for 2 of the measured anti-SARS-CoV-2 monoclonal antibodies (IgM and IgG). However, anti-RBD IgA concentrations were detectable in all samples and decayed rapidly (*τ*_*1*/*2*_ = 1.3 d, **Fig. 4B**).

It is becoming increasingly evident that a significant proportion of individuals with positive SARS-CoV-2 RT-qPCR test results remain asymptomatic or minimally symptomatic [5, 17-20] yet still contagious [5, 20]. However, ours is the first report on longitudinal analysis of an asymptomatic individual who maintained low SARS-CoV-2 RNA copy numbers and did not appear to trigger a traditional host immune response. This result is not entirely unexpected as antibody responses are not detectable in all COVID-19 patients, especially those who experience low grade symptoms [17, 21].

One household member (subject 39) was symptomatic and clinically diagnosed with COVID-19 by RT-qPCR and began participating in the study during convalescence after symptoms resolved (**Fig. 3A**). The subject developed a robust immune response in terms of anti-SARS-CoV-2 IgM, IgG, and IgA serum concentrations (**Table 3, Fig. 4C**) that persisted for over 2.5 months from the onset of symptoms. While the IgM and IgA concentrations decreased ca. 5-fold between April 14 and May 26, the IgG concentrations remained stable over the studied 2.5 months (**Fig. 4C**). This individual continues to be followed to measure the serum IgG kinetics, including concentration decay half-life and eventual background concentrations. The intense longitudinal RNA sampling provided a rare insight into the viral dynamics of an individual recovering from COVID-19 over a 3-month timeframe. Subject 39 remained positive for viral RNA for at least 71 days, one of the longest periods of SARS-CoV-2 shedding reported to date. Little is known about this potential subpopulation of individuals with persistent, long-term viral recalcitrance.

Intermittent news reports suggest that some convalescing COVID-19 patients who have tested negative for SARS-CoV-2 RNA by RT-qPCR may retest positive later in recovery, prompting speculation that some individuals are vulnerable to reinfection. Others hypothesize that positive retests are the result of non-infectious, residual viral fragments. Unfortunately, many of these reports are anecdotal and not scientifically controlled. Scientific studies on the alleged SARS-CoV-2 reinfection phenomenon are scarce. There are isolated case reports where a single COVID-19 patient was discharged, in part because of two consecutive negative SARS-CoV-2 RT-qPCR clinical test results, but re-tested positive for SARS-CoV-2 RNA during convalescence [22-26], including a 71-year old woman who tested positive for 60 days from the onset of symptoms (55 days from her first positive test) [27]. Other accounts detail similar results for small cohorts of 2 or more individuals [28, 29]. In a clinical study involving 66 patients who recovered from COVID-19, 11 (16.7%) retested positive for viral RNA in stool samples during convalescence [30]. A cohort of 86 COVID-19 patients was retested by RT-qPCR less than 28 days after self-reported symptom resolution. Eleven subjects (13%) were still diagnosed as positive by RT-qPCR at a median of 19 days (range 12-24 days). An *et al*. followed 262 COVID-19 patients for 2 weeks following discharge and found that 14.5% tested positive for SARS-CoV-2 RNA, suggesting that this subset may be carriers of the virus [31]. In a similar study, Huang *et al*. found that out of 414 recovering COVID-19 patients, discharged and subsequently quarantined, 69 (16.7%) retested positive for SARS-CoV-2: 13 with 2 readmissions, and 3 with 3 readmissions [32], suggesting that in some cases the virus was replication competent.

All Oak Crest employees with the exception of subject 31 remained negative for SARS-CoV-2 RNA over the course of the 3-month study. In our hands, the RT-qPCR test appeared to provide reliable results, with no known false positives or negatives. With the exception of subjects 31 and 39, inconclusive measurements (i.e., only one of the two oligonucleotide probes targeting the viral nucleocapsid protein gene transcript fragment met the assay threshold for positivity) made up a small fraction (0.32%) of the test results. We contend that in the context of a person developing or recovering from COVID-19 infection, inconclusive SARS-CoV-2 RNA measurements can define the transition phase between positive and negative, as supported by **Fig. 3**. Keeping in mind data published regarding the potential re-infectivity of SARS-CoV-2, subjects 31 and 39 were continuously retested, even following occasions where subjects received 2 consecutive negative results. Scattered inconclusive results accompanying days of high *Ct* values for subject 31 and 39 provide insight into the potential for re-infection and speak to the sensitivity of our assay. This study is especially useful in distinguishing between cases that appear to be true re-infections and those that could be classified by the transition from a positive to a negative result.

Unexpectedly, 9 of 16 employees (56%) tested positive for anti-SARS-CoV-2 antibodies, mostly IgM, at baseline between March 27 and April 3 (**Table 3**) suggesting possible exposure to the virus prior to the start of the clinical study. Many of these participants reported suffering from symptoms consistent with COVID-19 –thought to be a virulent flu strain at the time– in the first half of February. However, none of these serum samples had detectable concentrations of IgG, which has the longest half-life of the three measured virus-specific antibodies (**Fig. 4**) [33]. The findings remain unexplained and could be due to cross-reactivity in the IgM assay. Subject 18, a self-quarantined employee who had just recovered from suspected COVID-19 (based on symptomology) at the start of the study, repeatedly tested negative for SARS-CoV-2 RNA, but tested positive for IgM antibodies that rapidly declined (*τ*_*1*/*2*_ = 8.8 d, **Fig. 4A**). No IgG or IgA antibodies were detected in serum samples from this participant.

There are some caveats to interpreting our antibody data. We measured only IgM, IgG, and IgA against RBD and not the whole spike or E proteins, as discussed in more detail elsewhere [33]. The IgM assay is subject to greater uncertainty compared to IgA and IgG at low concentrations (ca. 0.25 µg mL^-1^, or lower) need to be interpreted with caution. However, all serum samples were analyzed at three levels of dilution (1:100, 1:40, and 1:20) and only detectable concentrations in at least 2 of the 3 diluted samples are reported.

The clinical study described here has met its goals to date by providing a safe work environment for employees of a small academic institute. The ability to perform routine, frequent COVID-19 testing not only has allowed us to responsibly maintain a productive work environment during the pandemic, but also provided continuous reassuring data for the participating households, many including vulnerable individuals. This had the important mental health benefits of reducing anxiety and providing a sense of normalcy in the workplace (i.e., safe zone) during an acutely stressful period.

Modeling local study impacts predicted that without our intervention up to 7 employees or household contacts could have become infected with SARS-CoV-2 by July 7, 2020. Our finding that only 2 subjects contracted COVID-19, one prior to the start of the study, suggests that workplace disease surveillance based on frequent, longitudinal testing combined with recommended public health practices (social distancing, frequent hand sanitizing, and mask wearing on select days), when feasible, can be effective at creating a safe zone or bubble preventing COVID-19 from entering into the workplace.

Our simulations predicted an important, local public health benefit from our scalable surveillance plan. Similar approaches are being adopted by professional sports leagues (e.g., the National Hockey League), the entertainment industry, and others to support responsible resumption of their activities. The study also generated important, new scientific data on the SARS-CoV-2 host dynamics enabled by its longitudinal, intense sampling design. Our clinical study represents a powerful example on how an innovative public health initiative can be dovetailed with scientific discovery.

## Data Availability

All relevant data are within the manuscript.
Restrictions apply to personal information on study participants.

## Acknowledgements

The study was funded entirely through discretional, institutional funds. The authors gratefully acknowledge this support and Oak Crest Institute of Science’s dedication to providing a safe work environment for its employees. We thank Alexandra de Jong, Susan Abramson, and the Aspire IRB staff for expediting the review of the study materials. We also gratefully acknowledge all study participants for donating their time and the valuable clinical specimens.

## Notes

### Competing Interest Statement

The authors have declared no competing interest.

### Funding Statement

The author(s) received no specific funding for this work

